# Lineage-Specific Transcriptional Response to Chikungunya ECSA and West African Lineages in Primary Human Chondrocytes

**DOI:** 10.64898/2026.03.10.26348026

**Authors:** Mario Rankin, Yashica Ganga, Victoria Sviridchik, Kajal Reedoy, Zesuliwe Jule, Adryele Figueiredo Pinheiro, Abdou Padane, Abed Nasereddin, Idit Shiff, Yuval Nevo, Inbar Plaschkes, Somasundram Pillay, Leonard Marais, Wesley C. Van Voorhis, Souleymane Mboup, Isadora C. de Siqueira, Khadija Khan, Alex Sigal

## Abstract

Chikungunya virus (CHIKV) causes long-term arthritis in about a third of infected people. It circulates as three main lineages. The East/Central/South African (ECSA) lineage and its derivatives have led to outbreaks in the Indian Ocean, South America, Pakistan, and most recently China, while the West African (WA) lineage is restricted to West Africa. Here we characterized CHIKV lineage-specific differences in human chondrocyte infection, the cartilage secreting cell type central to joint function. We compared transcriptional responses to ECSA and WA in chondrocytes isolated from eight donors undergoing clinically indicated orthopedic surgical procedures. We used serial viral dilutions so that each donor line included at least one ECSA and one WA infection with a similar proportion of infected cells. We performed RNA-Seq 24 hours post-infection. ECSA and WA induced substantially different transcriptional programs in chondrocytes. Compared to matched uninfected controls, we identified 1,235 genes significantly differentially regulated in ECSA and 842 in WA infection, with 604 overlapping. Infection with both lineages upregulated interferon response genes, and reduced expression of cell cycle-associated genes. However, only ECSA significantly reduced expression of NF-κB response genes, while WA infection strongly increased SOX2 expression, and uniquely increased NOTCH1 and reduced SOX9 expression, a pattern broadly consistent with a chondrocyte de-differentiation or loss of cartilage identity. These results show that CHIKV lineages elicit distinct transcriptional programs, with potential implications for lineage-specific joint pathology and disease course.

**Author Summary:** Chikungunya virus (CHIKV) infection often causes severe joint pain and arthritis that can persist for months or years. Different viral lineages of CHIKV circulate globally but the effect of viral lineage on the cellular responses in relevant infected cell types remains poorly understood. Here we infected primary human chondrocytes, the cells responsible for maintaining cartilage, with two major CHIKV lineages under matched infection conditions. Although both viruses replicated similarly and triggered strong antiviral interferon responses, they induced distinct transcriptional programs, with only ECSA suppressing the inflammatory NF-κB response and WA infection resulting in a chondrocyte gene expression pattern more consistent with de-differentiation. These lineage-specific effects on cartilage cell biology may contribute to differences in joint outcomes following infection.

## Introduction

Chikungunya virus (CHIKV) is a positive-sense RNA virus in the Alphavirus genus and a major cause of infection induced arthritis worldwide, as CHIKV infection may lead to persistent inflammatory joint disease in about one third of infected people [1–5]. CHIKV is an enveloped virus with a positive-sense RNA genome of 11.8 kb which encodes 9 proteins organized into two open reading frames. The virus is known to infect cells through the MXRA8 receptor [6] and uses the nonstructural protein nsP2 for transcription shutoff and translational modulation of host gene expression [7–10].

CHIKV has three primary lineages: ECSA, WA, and the Asian lineage. ECSA has evolved into several lineages/sub-lineages [1] including the Indian Ocean lineage (IOL), responsible for extensive infection in the Reunion Islands in 2006. It has also spread extensively worldwide, including to South America in 2012 and it is now endemic in Brazil and with epidemic spread in Paraguay [11]. Recently, it has led to epidemics in Pakistan and has been identified in China [12]. ECSA differs from WA in its ability to be spread by *Aedes albopictus* in addition to the *Aedes aegypti* mosquito due to a recent mutation in the envelope gene E1 [13].

CHIKV has been shown to infect cells in the joint including chondrocytes [2, 14]. Joints can become inflamed during infection and the resulting immune activation, and persistent inflammation of joint tissues leads to the hallmarks of arthritis [2, 5]. Joint surfaces consist of hyaline cartilage secreted by embedded chondrocytes. Hyaline cartilage reduces friction and absorbs mechanical stress. Joints also contain synovium and synovial fluid that is populated by synovial macrophages responsible for immune surveillance and homeostasis, as well as synovial fibroblasts responsible for maintaining the synovial environment [2, 15]. Chondrocytes are embedded in the cartilage they secrete which consists of type II collagens as well as aggrecan [16]. Chondrocytes differentiate from mesenchymal stem cells (MSCs) through a chondrogenic program orchestrated primarily by the transcription factor SOX9 [17]. *Ex-vivo* culture of primary human chondrocytes results in dedifferentiation back to MSCs [18].

Whether the infecting CHIKV lineage influences host cellular responses remain poorly defined. Since chondrocytes are the cellular component of articular cartilage, we reasoned that chondrocyte infection could contribute to cartilage damage and joint pathology. We therefore compared host transcriptional responses to the ECSA and WA CHIKV lineages in primary human chondrocytes under infection-matched conditions. We found that while ECSA and WA both elicit interferon and cell cycle arrest responses, there are substantial differences between the transcriptional response to each lineage, including down-modulation of the NfκB inflammatory response pathway which is significant with ECSA but not with WA infection.

## Results

### Chondrocyte isolation and validation of cell identity

We isolated and expanded joint cells from discarded surgical tissue from ten donors who underwent orthopedic surgery. Informed consent was obtained (Materials and Methods). Table 1 lists donor demographics and indications. After expansion, cells were frozen for future experiments (Materials and Methods). To examine the identity of the joint cell lines, we performed bulk RNA sequencing and used the gene expression pattern for reference-based cell type classification by SingleR with the Human Primary Cell Atlas [18].

**Table 1:** Chondrocyte donors in the study.

| PID | Age | Sex | Surgery type | Indication for surgery | Surgical site | HIV status |
| --- | --- | --- | --- | --- | --- | --- |
| 0001 | 60-70 | M | Joint replacement | Osteoarthritis | Wrist | + |
| 0004 | 30-40 | M | Joint replacement | Severe trauma | Elbow | + |
| 0008 | 30-40 | M | Joint replacement | Severe trauma | Elbow | - |
| 0012 | 0-10 | M | Arthrodesis | Foot deformity | Talar-navicular joint | - |
| 0014 | 50-60 | M | Joint replacement | Osteoarthritis | Hip | - |
| 0016 | 0-10 | F | Excision arthroplasty | Wrist deformity | Wrist | - |
| 0017 | 30-40 | M | Arthrodesis | Developmental foot deformity | Talar-navicular joint | - |
| 0022 | 30-40 | M | Joint replacement | Severe trauma | Elbow | - |
| 0024 | 40-50 | F | Excision arthroplasty | Delayed presentation radial head fracture | Elbow | + |
| 0033 | 40-50 | F | Excision arthroplasty | Severe trauma | Elbow | + |

All donor derived cell lines, with one exception, had a gene expression signature which most closely correlated with either chondrocytes or MSCs, the precursor cells for chondrocytes (Table S1). The MSC result is expected, as chondrocytes undergo dedifferentiation during *ex-vivo* culture [19]. The remaining donor derived cell line correlated most closely with a fibroblast gene expression signature (Table S1). This may be consistent with isolation and expansion of synovial fibroblasts present in the joint, or isolation of fibroblasts from fibrocartilage due to osteoarthritis or old trauma. This cell line was excluded from subsequent experiments. In addition, chondrocyte cells derived from donor 8 did not grow sufficiently well in culture and were also excluded to make experiments tractable. Experiments and transcriptome analysis were performed with the remaining 8 chondrocyte lineage cell lines.

To examine whether chondrocytes express the known CHIKV receptor MXRA8, we detected MXRA8 expression by flow cytometry in one of the chondrocyte lines as well as in the Vero-E6 cell line, known to be susceptible and permissive for CHIKV infection [20]. Both chondrocytes and Vero-E6 cells showed MXRA8 expression on a subset of cells with higher proportion of MXRA8 on the chondrocyte line relative to Vero-E6 cells (Fig. S1).

### ECSA and WA infections show separation by host gene expression

We isolated ECSA and WA viruses from plasma samples (Materials and Methods) from Brazil and Senegal. As expected, phylogenetic analysis showed the isolate from Brazil clusters with ECSA sequences, while the Senegal isolate clusters with WA sequences (Fig. S2). We used Vero-E6 cells to titer the CHIKV ECSA and WA isolates (Fig. S3) and infected chondrocyte lines and Vero-E6 cells with serial dilutions of ECSA and WA calibrated to result in comparable infection levels in chondrocytes for both lineages (Materials and Methods). Infections were performed for 24 hours in five independent experiments done on different days. Each experiment consisted of ECSA and WA infections (three dilutions) and three uninfected controls of 1 to 2 chondrocyte donors, as well as Vero-E6 cells infected with the same titer of ECSA or WA (Fig. S4–S8). Fig. 1A shows a representative chondrocyte line 24 hours post-infection. Interestingly, at the same viral titer, ECSA showed higher infection of chondrocytes relative to Vero-E6 cells. In contrast, WA infection was similar between chondrocytes and Vero-E6 cells (Fig. S9). At the 24-hour timepoint, RNA was extracted and used for bulk RNA-Seq.

**Figure 1:**
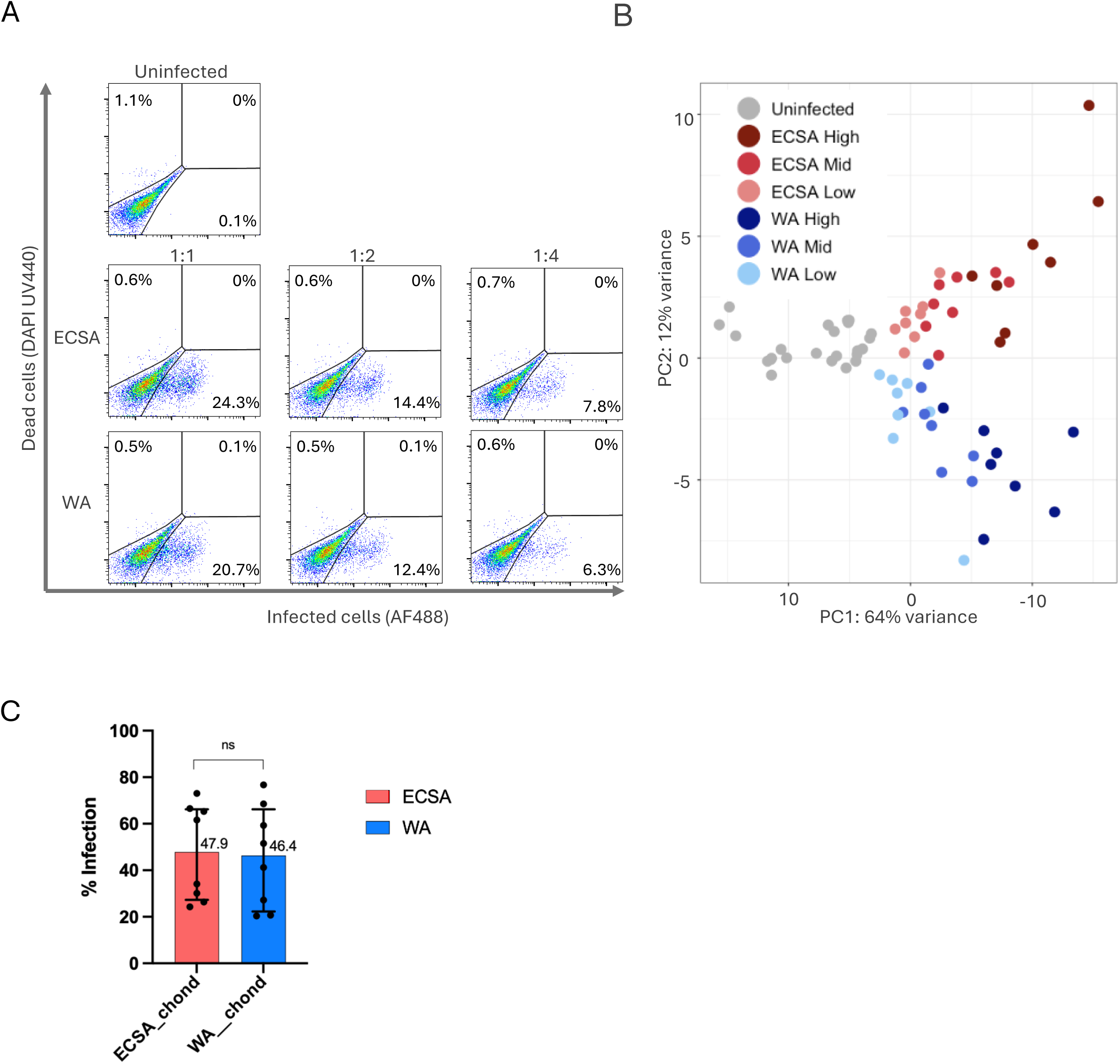
ECSA and WA infected chondrocytes show separation based on gene expression. (A) Flow cytometry plots of chondrocytes 24 hours post-*ex-vivo* infection with 2-fold serial dilutions of ECSA or WA. X-axis shows infection, y-axis shows live/dead stain, with percentage of live infected chondrocytes in the bottom right quadrant. Representative infection of chondrocytes from one donor (donor 0004), with other donors shown in Fig. S2. (B) Principal component analysis (PCA) of gene expression profiles for uninfected, ECSA-infected, and WA-infected chondrocytes derived from 8 donors. Each point represents one infected or uninfected primary chondrocyte line derived from one donor, with grey points denoting uninfected chondrocytes (3 repeats per donor) or infected chondrocytes (6 per donor, infected at one of three virus dilutions of ECSA or WA). (C) Median infection levels across chondrocyte donors where the closest matching infection condition in terms of proportion of infected cells was selected between ECSA and WA per donor. Numbers above bars represent the median values. Difference was not significant by the Wilcoxon test.

Principal component analysis of the RNA-Seq expression data revealed that principal component 1 (PC1) segregated samples by infection level, while PC2 separated ECSA- and WA-infected samples (Fig 1B). Genes in PC1 and PC2 included interferon response associated genes (Tables S2 and S3 show the top 10 leading genes in PC1 and PC2, respectively). Because of the effect of infection dose on the transcriptional response observed in PC1, our subsequent analyses were restricted to donor-matched samples where ECSA and WA infected a similar proportion of cells (Fig. S10). Median infection levels were 48% for ECSA and 46% for WA in this set of samples, a non-significant difference (Fig. 1C).

### Differentially expressed genes in ECSA and WA infection

We next examined which genes were differentially expressed in ECSA and WA infections relative to matched uninfected controls. We derived significantly differentially expressed (DE) genes in the infection matched samples by using FDR < 0.05 and absolute fold change > 1.5 relative to controls. Hierarchal clustering of the DE genes matrix per sample displays clustering of infected versus uninfected samples, as well as clustering of the WA infected samples (Fig. 2).

**Figure 2:**
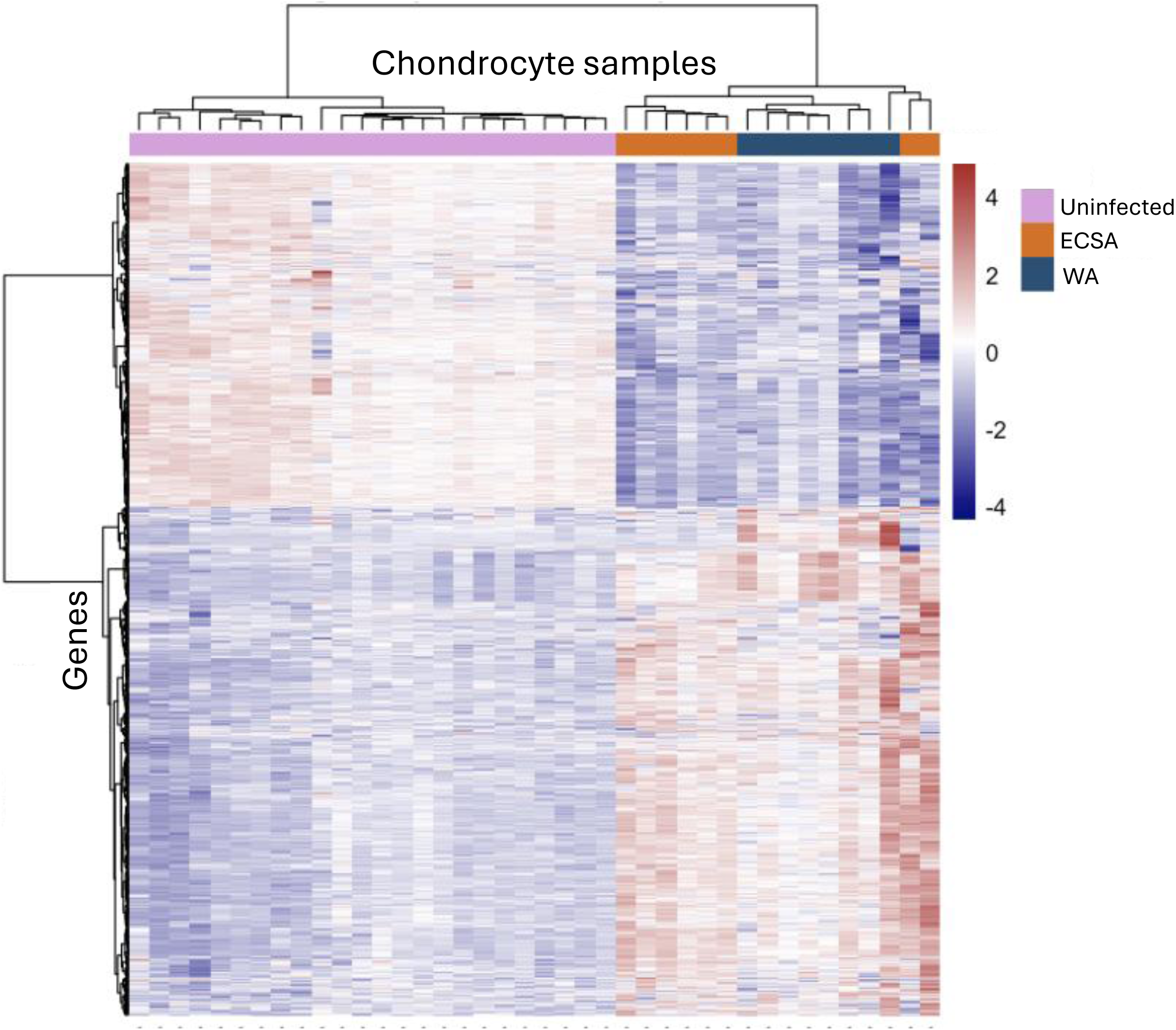
Heatmap of significantly differentially expressed genes in ECSA and WA infection. Donor matched samples where ECSA and WA infection differed by less than x% were compared to uninfected controls and significantly differentially expressed genes were determined as adjusted p < 0.05, Benjamini-Hochberg; |log₂ fold change| > 0.58 in either ECSA- or WA-infection.

We found that 1,235 DE genes were in ECSA infection and 842 in WA infection, with 604 (49%) overlapping, about half of the total (Fig. 3A). This included 756 (61%) upregulated and 479 (39%) downregulated DE genes in ECSA infection and 524 (62%) upregulated and 318 (38%) downregulated DE genes in WA infection. Of the 756 upregulated genes, 401 (53%) overlapped, while 203 of 604 (34%) of the downregulated genes overlapped, with ECSA showing higher numbers of non-overlapping genes both in the upregulated and downregulated subsets (Fig 3A). Both lineages exhibited markedly stronger upregulation (up to approximately 1,000-fold) relative downregulation (≤6-fold) (Fig. 3B). Many canonical interferon-stimulated genes involved in the cellular antiviral response were induced by both lineages, including IFIT1, MX1, OAS2, and OASL (Fig. 3B).

**Figure 3:**
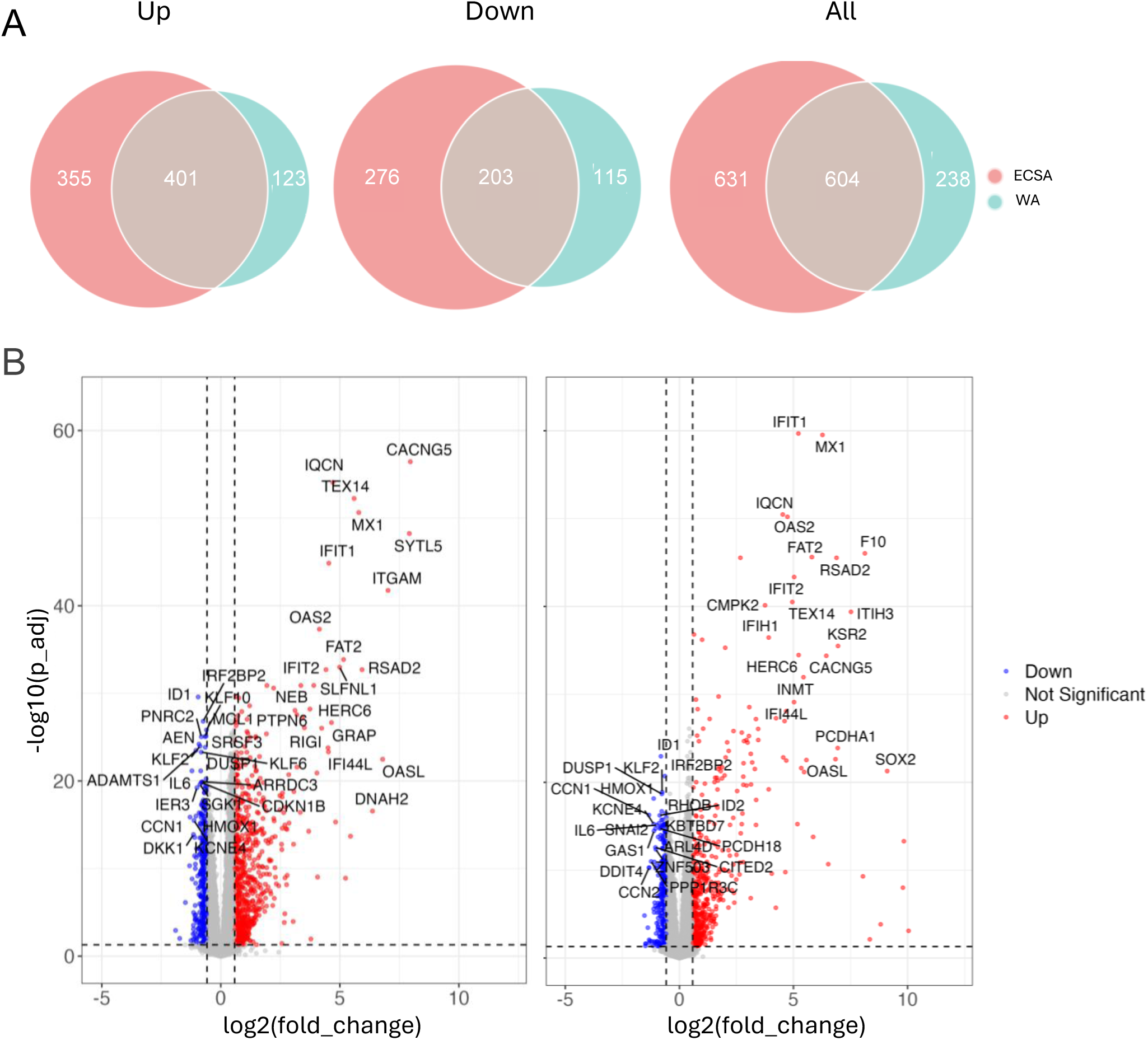
Overlap and distribution of differentially expressed genes in ECSA and WA infection. (a) Venn diagrams showing the overlap of significantly differentially expressed genes (adjusted p < 0.05, Benjamini-Hochberg; |log₂ fold change| > 0.58) between ECSA and WA infections compared to uninfected controls. Intersections are shown separately for upregulated genes (up), downregulated genes (middle), and all significant genes (down). (b) Volcano plots of differentially expressed genes in ECSA-infected (left) and WA-infected (right) chondrocytes compared to uninfected controls. Significantly upregulated (red) and downregulated (blue) genes are highlighted.

### Gene Set Enrichment Analysis shows unique pathways in ECSA infection

To examine cellular pathways induced by infection, we performed Gene Set Enrichment Analysis (GSEA, Materials and Methods). This demonstrated enrichment of interferon-α and interferon-γ response pathways in both lineages (Fig. 4A). Cell cycle–associated Hallmark gene sets, including E2F targets, were negatively enriched, indicating a cell cycle arrest response. Notably, the NF-κB Hallmark gene set was significantly negatively enriched only in ECSA infection (Fig. 4A).

**Figure 4:**
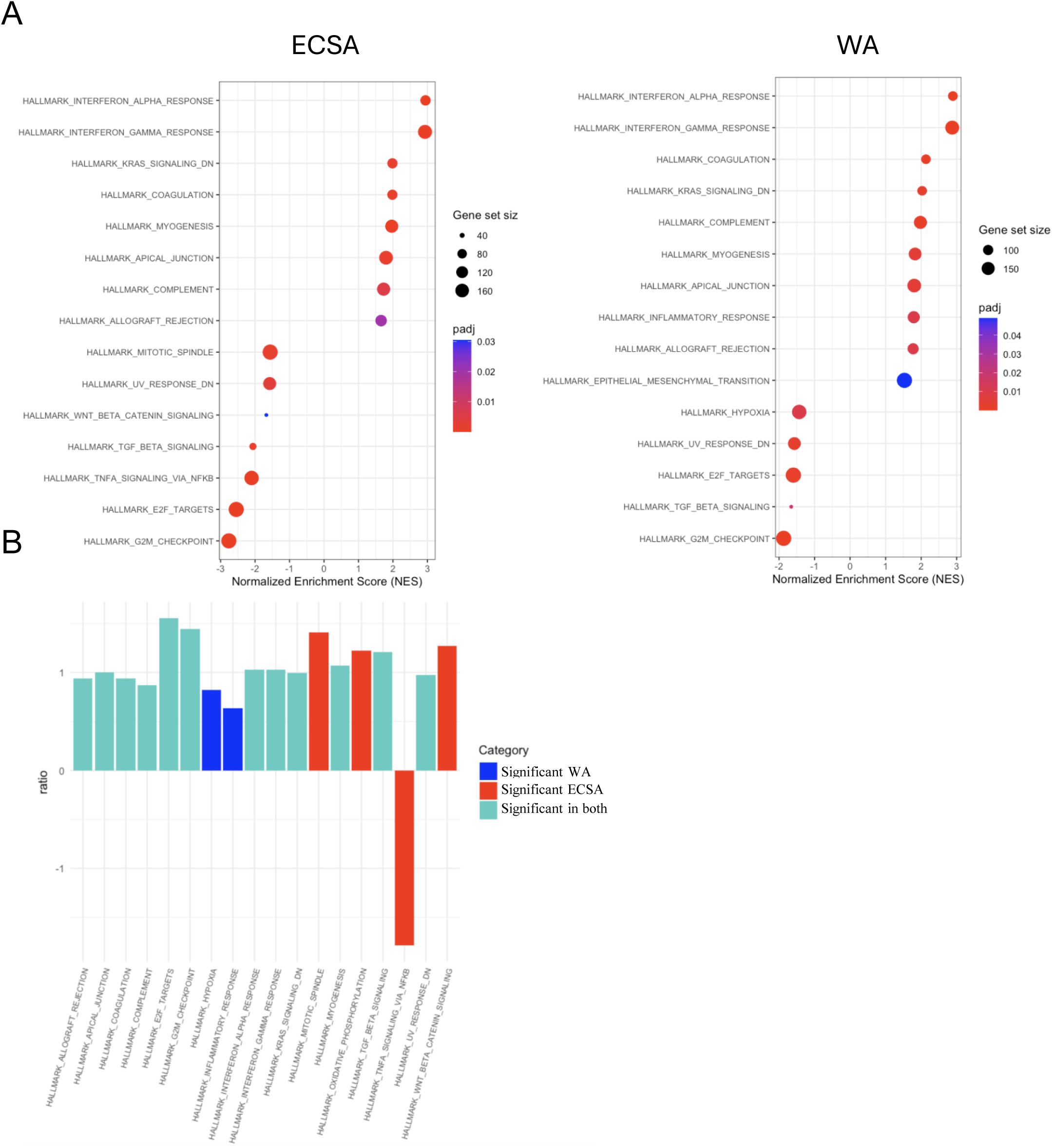
Gene ontology enrichment analysis of differentially expressed genes in ECSA and WA infection. (A) GO enrichment of significantly differentially expressed genes in ECSA-infected chondrocytes and WA-infected chondrocytes compared to uninfected controls (adjusted p < 0.05, Benjamini-Hochberg; |log₂ fold change| > 1.5). Dot size represents gene count and color indicates adjusted p-value. (B) Comparison of normalized enrichment scores (NES) for enriched pathways in ECSA and WA infection. Bar plot showing the ratio of NES for the union of significantly enriched pathways in ECSA-infected (n = 14) and WA-infected (n = 16) chondrocytes compared to uninfected controls. Pathways not reaching significance in one condition are indicated by red or blue color.

To compare differences in pathways between ECSA and WA, we divided the normalized enrichment score (NES) of ECSA by that for WA for all significantly regulated pathways (for a pathway to be included, it needed to be significant in either ECSA, or WA, or both). This gave a total of 18 pathways, where 12 were significantly regulated in both ECSA and WA, four were significant in ECSA only and two were significant in WA only (Fig 4B). All except for one pathway showed same directionality in ECSA and WA infection (i.e., both upregulated or both downregulated). The exception was the NF-κB pathway, which was only significantly downregulated in ECSA infection (Fig 4B).

### Chondrocyte-Specific Gene Regulation in ECSA

Lastly, we examined the effect of infection on chondrocyte-associated genes. We constructed a list of 80 previously known to be chondrocyte associated genes [17, 21–37] and examined whether they were significant DE genes in our dataset. ECSA infection induced significantly increased expression (FDR<0.05, fold change >1.5) of 15 chondrocyte-related genes, and decreased expression of 8 chondrocyte-related genes (Fig. 5A). Upregulated genes included the collagen COL2A1 and aggrecan (ACAN), key components of hyaline cartilage secreted by differentiated chondrocytes [38]. However, SOX2, one of four Yamanaka factors that when combined, are sufficient to reprogram differentiated cells to an embryonic-like state [35], was also upregulated. SOX2 may be involved in chondrocyte de-differentiation to progenitor cells [25, 37]. In addition, CCN1, CCN2, ID1, ID2, BMP2, and BMP4 involved in chondrocyte differentiation [30, 33, 34, 39], and HMOX1, which has been shown to be cytoprotective in chondrocytes by reducing oxidative stress [32], were all significantly reduced. CHRDL2, which is expressed in mesenchymal stem cells and is decreased upon differentiation to chondrocytes [31], was increased.

**Figure 5:**
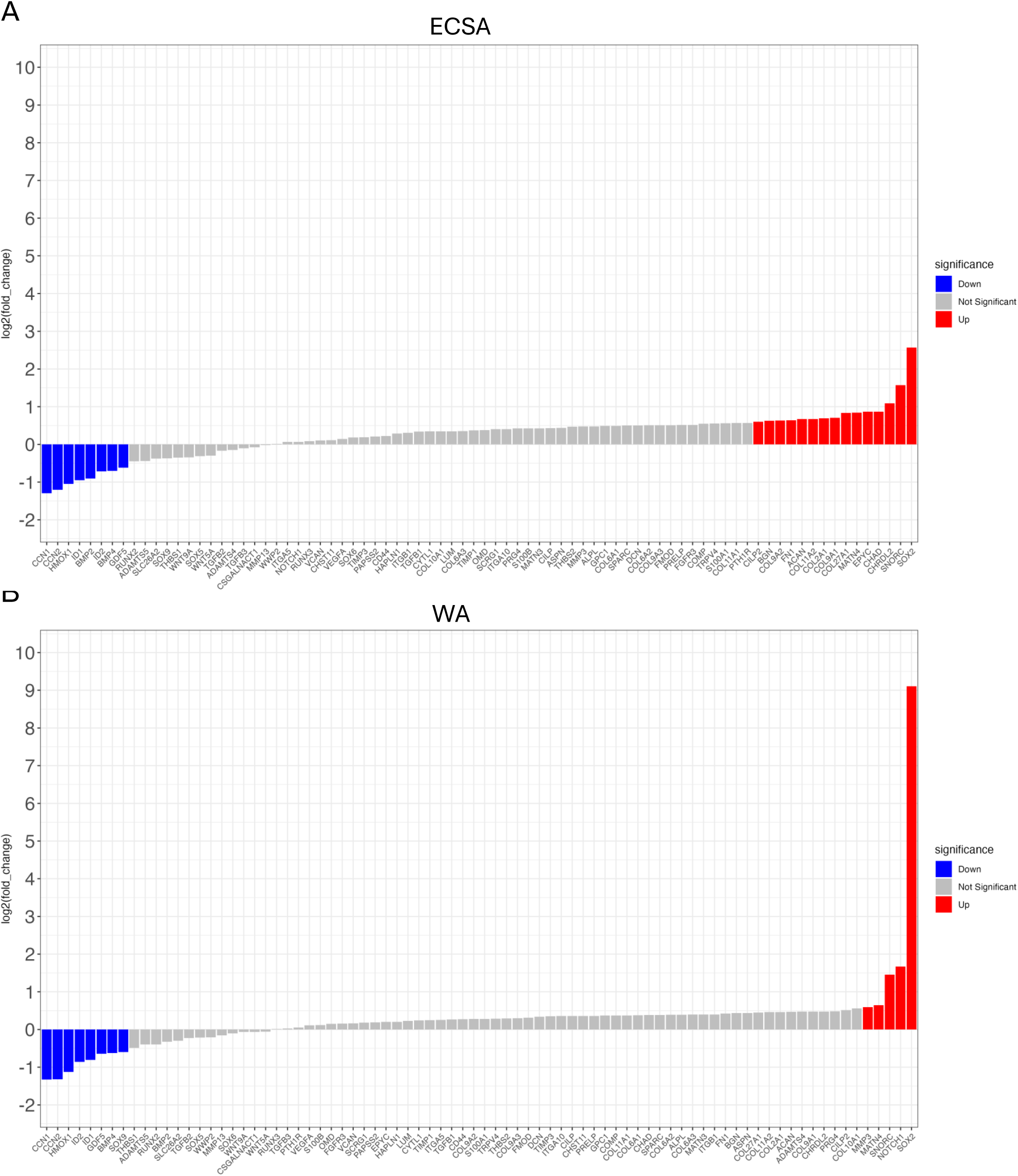
Expression of chondrocyte-associated genes in ECSA and WA infection. Bar plot showing log₂ fold change of chondrocyte-associated genes in ECSA (A)- and WA (B)-infected chondrocytes compared to uninfected controls. Significantly upregulated genes are shown in red, significantly downregulated in blue, and genes not reaching significance in grey.

While ECSA showed both increased and reduced expression of genes driving chondrocyte differentiation, WA infection induced a pattern mostly consistent with de-differentiation. ECSA infection induced significantly increased expression of 5 chondrocyte-related genes, and decreased expression of 8 chondrocyte-related genes. SOX2, involved in driving cells to a more de-differentiated state and which was upregulated in ECSA by about 5-fold, was upregulated in WA about 1000-fold (Fig 5B). Notch1, involved in chondrocyte homeostasis, but whose over-expression was shown to interfere with chondrocyte differentiation through decreasing SOX9 [28, 29], was significantly increased by WA but not ECSA infection. As in ECSA infection, CCN1, CCN2, ID1, ID2, BMP4 and HMOX1 were all significantly reduced. In addition, SOX9, the master regulator of the chondrocyte cell fate program which activates key cartilage genes such as COL2A1 and ACAN [17, 24], was also significantly downregulated in WA infection. In contrast to ECSA, COL2A1 and ACAN were not significantly upregulated with WA. Therefore, our findings suggest that WA infection shows stronger remodeling of the chondrocyte transcriptome to a de-differentiated state relative to ECSA infection.

## Discussion

We investigated CHIKV ECSA and WA lineage infection of primary chondrocytes, a readily infectable cell type expressing the MXRA8 receptor used by the virus. We observed that infection by CHIKV ECSA and WA lineages induced broadly similar interferon antiviral and cell cycle arrest responses in primary human chondrocyte cultures. In both lineages, upregulated genes had substantially stronger responses compared to downregulated genes. However, there were many genes that were uniquely up or downregulated in ECSA or WA.

The lineage-specific transcriptional differences involved a larger number of differentially regulated genes and significant suppression of the NF-κB signaling pathway with ECSA but not WA infection. In agreement with our results, a previous study showed that CHIKV nsP2 interfered with melanoma differentiation-associated protein 5 (MDA5)-mediated induction of the NF-κB pathway [8]. In addition, we found that WA infection showed a stronger signal for driving the chondrocyte transcriptome to a de-differentiated state. Relative to ECSA, WA infection resulted in about 200-fold higher upregulation of SOX2 expression, significant NOTCH1 upregulation, and a significant downregulation of SOX9. These differences could contribute to divergent joint outcomes following infection, as SOX9, SOX2, and NOTCH1 are all involved in the progression of arthritis [17, 24, 25, 27–29, 36, 37, 40, 41].

Limitations of the study include that *ex vivo* chondrocyte cultures may not fully reflect responses of differentiated chondrocytes in the joint tissue. Furthermore, the effects of ECSA and WA infection may differ in other CHIKV infectable cell types. The outcome of infection *in vivo* may also have additional dimensions, including initiation of the adaptive immune response and T cell infiltration [2] which may play a determining role in CHIKV infection outcome including long-term arthritis. Lastly, nsP2 of CHIKV and other alphaviruses is known to modulate translation of host genes [42, 43] as well as by other pathways [10, 42]. Hence CHIKV infection may further remodel cellular pathways, beyond the transcriptional response examined here.

This study is a step in the understanding of differences in ECSA and WA infection in terms of the response of a key cellular component in the joint. Both ECSA and WA are thought to cause long-term arthritis and evidence that they can lead to different outcomes is limited but does include a study in animal models showing differences in pathology, with more severe disease in WA infection [44]. Our observation that WA does not significantly suppress NFκB may indicate that WA may cause a stronger inflammatory response and therefore lead to higher pathology [45].

Our results showing gene markers of chondrocyte de-differentiation in CHIKV infection, which are most pronounced in the WA lineage, may indicate that CHIKV may damage chondrocyte function in cartilage by transcriptional reprogramming. While infected cells are unlikely to survive long either because of the cytotoxic effects of infection or the adaptive immune response to it, long-term effects are possible if bystander cells are also reprogrammed. In addition, these results suggest the possibility that not only host risk factors such as pre-existing joint pathology predispose CHIKV infected individuals to long-term arthritis [46], but that different strains of CHIKV may also induce different infection outcomes.

## Materials and Methods

### Ethics statement

Primary human chondrocytes were collected through a prospective study approved by the Biomedical Research Ethics Committee at the University of KwaZulu-Natal (reference BREC/00005351/2023). Consent was obtained from patients undergoing clinically indicated surgical procedures. At the time of surgical consent, research consent was obtained for collection of hyaline cartilage tissue samples, that would have otherwise been discarded and incinerated post-surgical procedure. These surgical procedures include amputations, prosthetic joint replacements and joint arthrodesis. The ECSA lineage was isolated from clinical samples obtained from an ongoing arbovirus surveillance study approved by the IGM-Fiocruz IRB (CAAE 53378421.7.0000.0040). Written informed consent was obtained from patients presenting with acute febrile illness within the previous seven days, accompanied by at least one additional symptom, including myalgia, arthralgia, nausea, or rash at the Emergency Care Unit UPA Santo Antonio, in Salvador, Brazil, between March and July 2023. The WA lineage was isolated from participants enrolled to participate in a study approved by the Comite National d’Ethique pour la Recherche au Senegal (CNERS) (Ref. Number 000031/MSAS/CNERS/SP/06/02/2023). Samples were collected with informed consent from participants in the Kedougou district during the 2023 Chikungunya outbreak. The Direction de la Prevention, part of the Senegal Ministry of Health, was informed of the IRESSEF intervention during the chikungunya outbreak in Senegal on August 8, 2023 (N/Ref: SM/IRESSEF/RARS/116.09.2023).

### Reagent availability statement

Viral isolates are available upon reasonable request. Sequences of isolated ECSA and WA lineages used in this study have been deposited in GISAID with accession numbers as follows:

| **Virus** | **GISAID** |
| --- | --- |
| **CHIKV ARB18 (ECSA)** | EPI_ISL_19458433 |
| **CHIKV ARB113 (WA)** | EPI_ISL_20146493 |

### Vero-E6 cells

Vero-E6 cells overexpressing TMPRSS2, originally from BEI Resources (NR-54970) were used for experiments. The Vero-E6 cell line was propagated in growth medium consisting of Dulbecco’s Modified Eagle Medium (DMEM, Gibco 41965-039) with 10% fetal bovine serum (Hyclone, SV30160.03) containing 10mM of hydroxyethylpiperazine ethanesulfonic acid (HEPES, Lonza, 17-737E), 1mM sodium pyruvate (Gibco, 11360-039), 2mM L-glutamine (Lonza BE17-605E) and 0.1mM nonessential amino acids (Lonza 13-114E).

### Chondrocyte cell isolation from hyaline cartilage

A specimen size of approximately 5mm x 5mm x 1mm of hyaline cartilage was dissected into smaller pieces (approximately 1-2mm^3^ each) either with a sharp blade dissection or with a bone nibbling rongeur instrument. The specimen fragments were then transferred into a sterile container containing 5mL chondrocyte medium ((Dulbecco’s Modified Eagle Medium (DMEM, Gibco 41965-039) with 10% fetal bovine serum (Hyclone, SV30160.03) containing 10mM of hydroxyethylpiperazine ethanesulfonic acid (HEPES, Lonza, 17-737E), 1mM sodium pyruvate (Gibco, 11360-039), 2mM L-glutamine (Lonza BE17-605E), 0.1mM nonessential amino acids (Lonza 13-114E) and 1 x Insulin-Transferrin-Selenium (ITS-G) (Capricorn – ITS-H)) and processed within an 1 h after tissue collection. Cartilage tissue was rinsed in phosphate-buffered saline (PBS) (Capricon, PBS1-A) with 2.5 µg/mL Fungizone (Gibco, 15290018). Tissue was digested enzymatically with 0.25% Trypsin-EDTA (Thermo Fisher Scientific – 25200072) for 15mins at 37°C and centrifuged at 300 RCF for 5 mins. For the main enzymatic digestion, tissue was added to 0.7 mg/mL collagenase type II (Thermo Fisher Scientific, 17101015) in chondrocyte medium and digested for 4 h at 37°C with shaking at 300 rpm. The cell suspension was filtered through a 70 µM strainer (Falcon, BD352350), washed twice with chondrocyte medium, resuspended in chondrocyte medium and added to a single well of a 24-well plate (TPP, Z707805).

### Chondrocyte cell propagation

Cells were propagated in chondrocyte medium, with a medium change at 24 hours and subsequently every 3-4 days. At 95% confluence (usually 1-2 weeks), cells were detached using 0.25% trypsin-EDTA and passaged into a larger well/vessel for expansion. Passage 1 was transferred from a well of the 24-well plate to a well of a 12-well plate (TPP, Z707775), passage 2 was transferred to a well of a 6-well plate (TPP, Z707759), passage 3 was transferred to a T25 flask (TPP, Z707481), passage 4 was transferred to a T75 flask (TPP, Z707546), passage 5 was expanded to 3 x T75 flasks and passage 6 was expanded to 9-10 T75 flasks. Once cells reached confluence in T75 flasks, they were detached using 0.25% Trypsin-EDTA, resuspended in 90% fetal bovine serum with 10% dimethyl sulfoxide (Sigma, D8418-50 mL) at 1-1.5 x 10^6^ cells per cryovial, and cryopreserved in liquid nitrogen.

### Isolation and expansion of ECSA and WA viral lineages

Work was performed in Biosafety Level 3 containment using protocols for CHIKV approved by the Africa Health Research Institute Biosafety Committee. For the isolation of the ECSA lineage, Vero-E6 cells were seeded at 8 × 10^4^ cells in a 24 well plate and incubated for 18–20 hours pre-infection. After one Dulbecco’s phosphate-buffered saline (DPBS) wash, the sub-confluent cell monolayer was inoculated with 100μL plasma diluted 1:2 with growth medium filtered through a 0.45μm filter (total 200μL). Cells were incubated for 2 hours at 37 °C, 5% CO_2_. Wells were then filled with 0.8mL complete growth medium supplemented with 10% DMSO and incubated at 37 °C, 5% CO2. After 4 days of infection (completion of passage 1 (P1)), supernatant was collected, cells were trypsinized, centrifuged at 300 RCF for 3 min and resuspended in 3mL growth medium. All infected cells and supernatant were added to VeroE6 cells seeded at 1.5× 10^5^ cells per mL, 5mL total, 18–20 hour earlier in a T25 flask for cell-to-cell infection. The coculture was incubated for 2 h and the flask was filled with 5mL of complete growth medium and incubated at 37 °C, 5% CO_2_. After 3 days of infection, (completion of passage 2 (P2)), supernatant was collected, cells were trypsinized, centrifuged at 300 RCF for 3 min and resuspended in 5mL growth medium. All infected cells and supernatant were added to Vero-E6 cells seeded at 1 × 10^5^ cells per mL, 20mL total, 18–20 hour earlier in a T75 flask for cell-to-cell infection. The coculture was incubated for 2 h and the flask was filled with 20mL of complete growth medium and incubated for 2 days at 37 °C, 5% CO_2_. The viral supernatant from this culture (passage 3 (P3) stock) was used for experiments. Isolation of the WA lineage was performed similarly, starting in a 6-well plate with a seeding density of 3 × 10^5^ per well in a total volume of 3mL. 4 days post infection, cells and supernatant were moved to a T25 flask. After 3 days, cells and supernatant were moved to a T75 flask and the virus supernatant was harvested after 2 days of infection.

### Infection and RNA isolation

Chondrocytes were calibrated with different viral dilutions of ECSA and WA to find dilutions where the percentage of ECSA and WA infected cells was comparable (data not shown). We found that ECSA at a multiplicity of infection (MOI) of 0.01 gave similar infection in chondrocyte lines to WA infection at an MOI of 0.05 at 24 hours post-infection, where MOI was measured in Vero-E6 cells. For infection experiments used for RNA-Seq, chondrocyte cell lines and Vero-E6 cells were plated at 1 × 10^5^ cells per well in a 12-well plate (TPP, Z707775) 1-day pre-infection. The following day, cells were infected with ECSA virus at an MOI of 0.01, 0.005, and 0.0025 and WA virus at an MOI of 0.05, 0.025, and 0.013 in 100 μL growth media per well. Cell-virus mixtures were incubated for 3 h at 37 °C, 5% CO_2_ followed by 3 washes with DPBS and replenished with 1 mL growth media. At 24 hours post-infection, cells were trypsinized, collected and stained with Blue Live/Dead stain as per manufacturer instructions (ThermoScientific, L34961). The samples were then washed in 1 mL PBS and resuspended in Cytofix/Cytoperm buffer (BD Biosciences, BD 554655) for 20 min at 4⁰C in the dark. The samples were then stained with 2 μg/mL monoclonal mouse anti–chikungunya virus antibody (L S Bio, LS–C203508) diluted in Perm/Wash buffer (BD Biosciences, BD 554723) and incubated 1 hour at 4⁰C in the dark. Samples were washed twice using Perm/Wash buffer (BD Biosciences), stained with 2 μg/mL goat-anti mouse AF488 IgG (BioLegend, 405319) diluted in Perm/Wash buffer and incubated 1 hour at 4⁰C in the dark. Cells were analyzed on an BD FACSymphony. Data was analyzed using FlowJo V10.10.0 and Graphpad Prism V10 software. For RNA isolation, total RNA was isolated using the RNeasy Mini Kit (Qiagen, 74104) as per manufacturer’s instructions. The protocol selected was the Purification of Total RNA from Animal Cells Using Spin Technology. Cells were washed three times with PBS and Buffer RLT was added. The homogenized cell lysates were collected and stored at -80 °C and later processed. On-column DNase digestion was performed using the RNase-Free DNase Set (Qiagen, 79254). RNA was quantified on the TapeStation 4200 using RNA ScreenTape Analysis (Agilant Technologies, 5067-5576). Agilent TapeStation Software v5.2 was used to generate RNA Integrity Number equivalent (RINe) values to assess RNA quality and integrity. RNA samples with RINe >7 were processed for RNAseq analysis.

### MXRA8 staining

Chondrocytes from donor 0004 and Vero-E6 cells were detached using 200-400 U/mL Accutase solution (Sigma, A6964) for 5-10 mins at room temperature, after the addition of 8 mL growth medium, cells were passed through a 40 μM cell strainer (Corning, 352340) and centrifuged at 300 x g for 5 mins. Purified NA/LE Human BD Fc Block (BD, 564765) was diluted in non-perm buffer [(0.5 g Bovine Serum Albumin (Rosche, 03117332001), 0.5 mL heat inactivated fetal bovine serum (Hyclone, SV30160.03) add 200 μL of 0.5 M EDTA (Sigma, 324506), 50 mL PBS (Capricon, PBS1-A)], added at a concentration of 2.5 μg/mL to 1 x 10^^5^ cells and incubated for 30 mins on wet ice. Cells were washed twice using non-perm buffer and stained with Blue Live/Dead stain as per manufacturer instructions (ThermoScientific, L34961). The samples were then stained with anti-MXRA8 antibody (L S Bio, LS–C203508) at 2 μg/μL diluted in non-perm buffer and incubated 1 hour on wet ice in the dark. Samples were washed twice with non-perm buffer, stained with 2 μg/mL goat-anti mouse PE (BioLegend, 405307) diluted in non-perm buffer and incubated 1 hour on wet ice in the dark. Cells were analysed on an BD FACSymphony. Data was analysed using FlowJo V10.10.0 and Graphpad Prism V10 software.

### Gene expression using RNA-Seq

RNA quality and quantity were determined by Agilent TapeStation analysis (Agilent). Stranded mRNA libraries were generated using the Watchmaker mRNA Library Prep Kit (Watchmaker Genomics 5744 Central Ave. Suite 100 Boulder, CO 80301) according to manufacture recommendations. In brief, RNA samples contained 1000-2000 ng in 50 ul of RNAse/DNAse free water were used for the library construction. Samples were heated for 4 min at 85 °C to achieve fragment library sizes for sequencing 300-400 bp. The KAPA Unique Dual-Indexed Adapter Kit (15 μM) kit (Roche, 08861919702) was used to assign a unique barcode to each sample through a ligation reaction, followed by bead purification and an enrichment step according to the manufacturer’s recommendations. Finally, libraries were eluted in 35 μl of elution buffer. Then they were adjusted to 10 nM pool, the adjusted pool was loaded into NovaSeqX machine (Illumina), as 161 bp paired end read. Binary Base Call (BCL) output files were converted to FASTQ format, using BCL to FASTQ (bcl2fastq v2.20.0.422 Copyright (c) 2007–2017 Illumina, Inc.). Library preparation and sequencing were performed at the Core Facility of the Hebrew University Faculty of Medicine.

### Differential expression analysis

Raw sequencing reads were quality-trimmed with cutadapt (v1.18), using a quality threshold of 32 for both ends, poly-G,A,T,C sequences and adapter sequences were removed, reads shorter that 20 bp were removed. The processed reads were aligned to a joined genome that included the host genome Homo sapiens version GRCh38and Chikungunya ECSA virus: NC_004162.2. Mapping was performed with STAR aligner (V.2.7.10a) while utilizing the relatively permissive alignment parameters: outFilterMismatchNmax 999, outFilterMismatchNoverLmax 0.33, outFilterScoreMinOverLread 0.33, outFilterMatchNminOverLread 0.33. This allowed reads which were derived from both WA and ECSA lineages to map to the one reference genome which was used. Genome annotations of the human genome were from Ensembl release 113. Quantification was done with htseq-count (V2.6.1). Only protein-coding genes were retained for downstream analysis. Ensembl gene identifiers were mapped to HGNC gene symbols using the org.Hs.eg.db R/Bioconductor annotation package. Count data were imported into R version 4.4.2 and analyzed using DESeq2 (v1.46) [47]. Analysis of differential gene expression was performed using DESeq2 R package where raw counts were normalized using the median of ratios default method of DESeq2 to adjust for sequencing depth and library composition. Differentially expressed genes were identified by a design formula accounting for donor effects and group, where group encoded the combination of CHIKV strain (ECSA or WA) and viral dilution, and donor accounted for inter-individual variability. Genes with low counts (<10 reads in at least 5 samples) were filtered out prior to analysis. Differentially expressed (DE) genes were identified using a threshold of | fold change| > 1.5 and adjusted p-value < 0.05 (Benjamini–Hochberg correction).

### Chondrocyte identity verification

Cell type identity was independently verified using SingleR (v2.8.0), a reference-based cell type classification method. The Human Primary Cell Atlas, comprising 713 microarray samples across 37 primary human cell types, was used as the reference dataset via the celldex R package (v1.16). Gene symbols were harmonized between test and reference datasets using the alias2SymbolTable function from limma (3.62.2). Each sample was assigned the cell type label with the highest correlation score, with fine-tuning enabled to improve discrimination between closely related cell types.

### Principal component analysis

Count data were transformed using the variance-stabilizing transformation (VST) implemented in DESeq2. To account for donor-specific effects, batch correction was applied to the VST-transformed matrix using the removeBatchEffect function from the limma package [48]. Principal component analysis (PCA) was performed on the corrected expression matrix and visualized using plotPCA.

### Heatmap generation

Heatmaps were generated using pheatmap to visualize the union of significantly differentially expressed genes identified in both ECSA vs. control and WA vs. control comparisons (n = 1,474). Row-scaled VST-normalized expression values were displayed using a diverging navy-to-white-to-firebrick color scale, with hierarchical clustering performed using Ward’s D2 method.

### Volcano plots

Volcano plots were generated using ggplot2, displaying log2 fold change versus −log10 adjusted p-value for all genes. Dashed lines indicate significance thresholds of |log2FC| > 0.58 and adjusted p-value < 0.05. Upregulated and downregulated genes are highlighted in red and blue, respectively, with selected genes labeled using ggrepel.

### Gene set enrichment analysis

Gene set enrichment analysis (GSEA) was performed on ranked gene lists for both CHIKV strains using the Hallmark gene set collection from the Molecular Signatures Database (MSigDB) (v25.1.1) [49]via the msigdbr and clusterProfiler (v4.14.6) R packages [50]. To identify shared and strain-specific transcriptomic responses, Venn diagrams were constructed comparing significantly upregulated, downregulated, and all significant DEGs between the ECSA and WA lineages. Expression patterns of overlapping DEGs were visualized as heatmaps. All analyses were performed in R (v4.4.2).

## Data Availability

All data produced in the present study are available upon reasonable request to the authors

---

**Figure S1:**
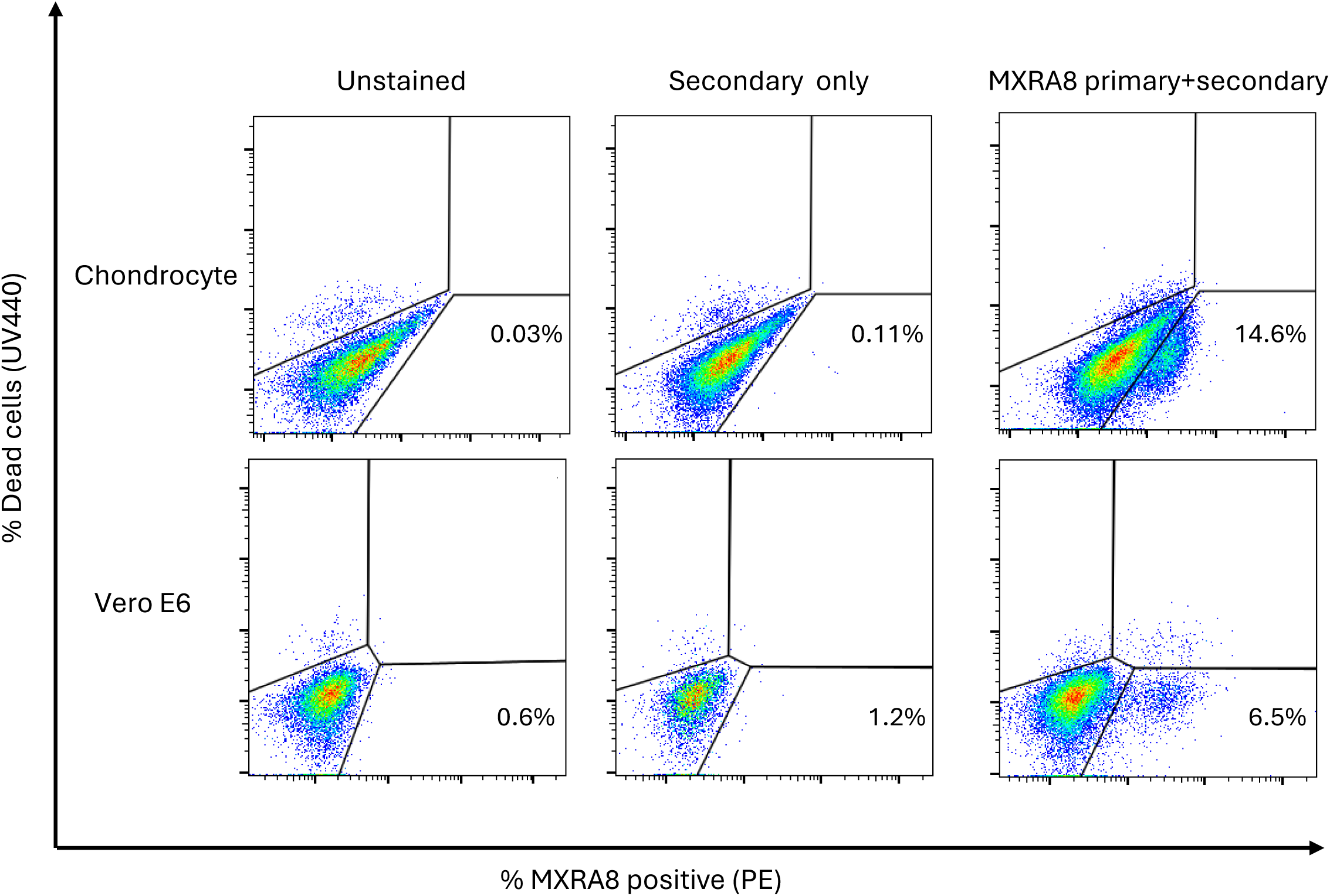
Surface expression of MXRA8 on chondrocytes and Vero-E6 cells. Flow cytometry dot plots of MXRA8 surface staining under non-permeabilized conditions. Top row: primary human chondrocytes (donor 0004). Bottom row: Vero-E6 cells.

**Figure S2:**
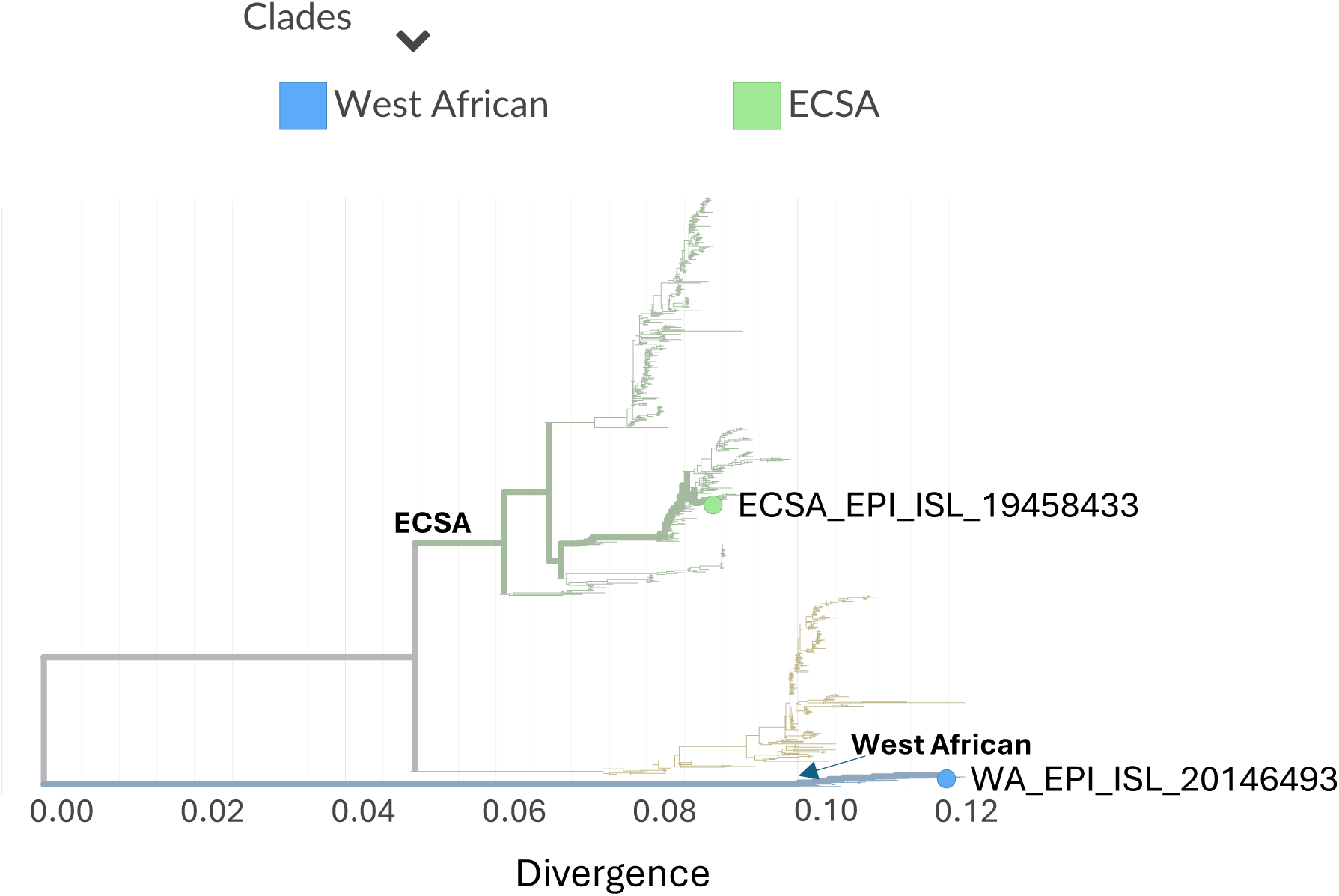
Phylogenetic analysis of CHIKV ECSA and WA isolates. Phylogenetic tree constructed using Nextclade, showing the evolutionary relationship between ECSA and WA lineage sequences isolated in this study. Branch lengths represent nucleotide substitutions per site.

**Figure S3:**
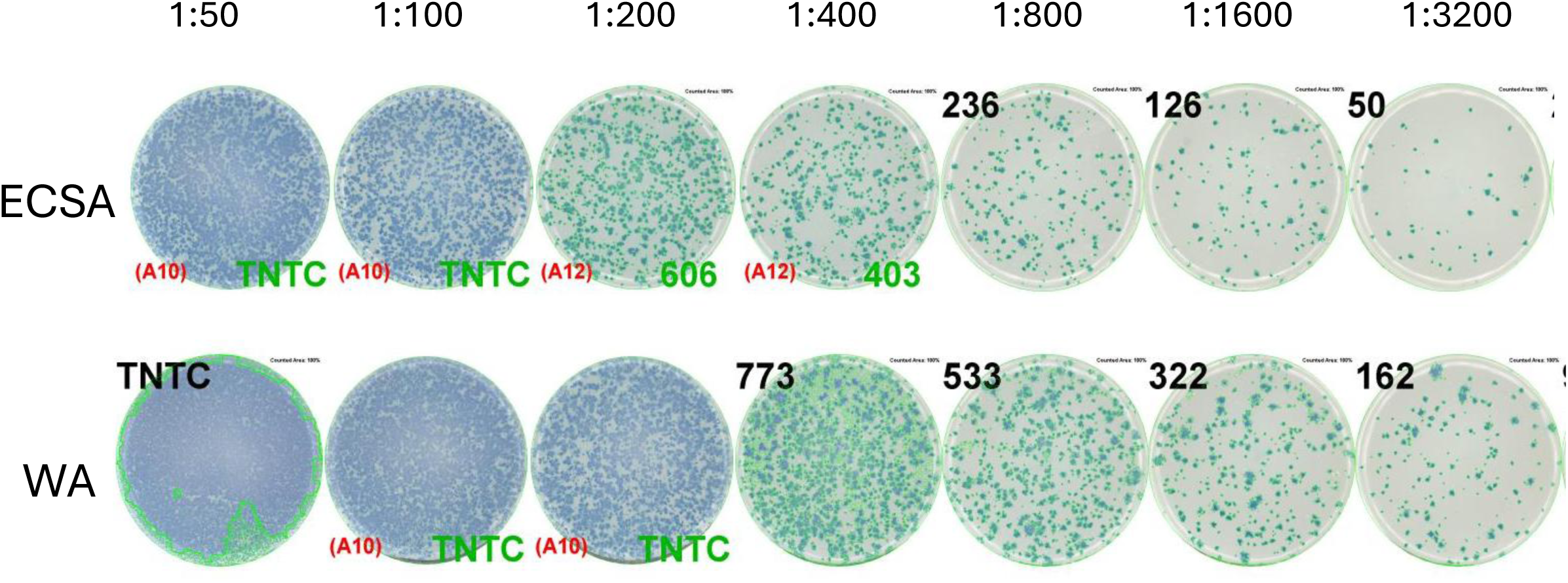
Titration of CHIKV ECSA and WA lineages on Vero cells. Viral titers determined by focus forming assay for ECSA and WA lineages across serial dilutions on Vero-E6 cells. Numbers represent focus-forming units (FFU) per well (100 uL input virus). TNTC: too numerous to count.

**Figure S4:**
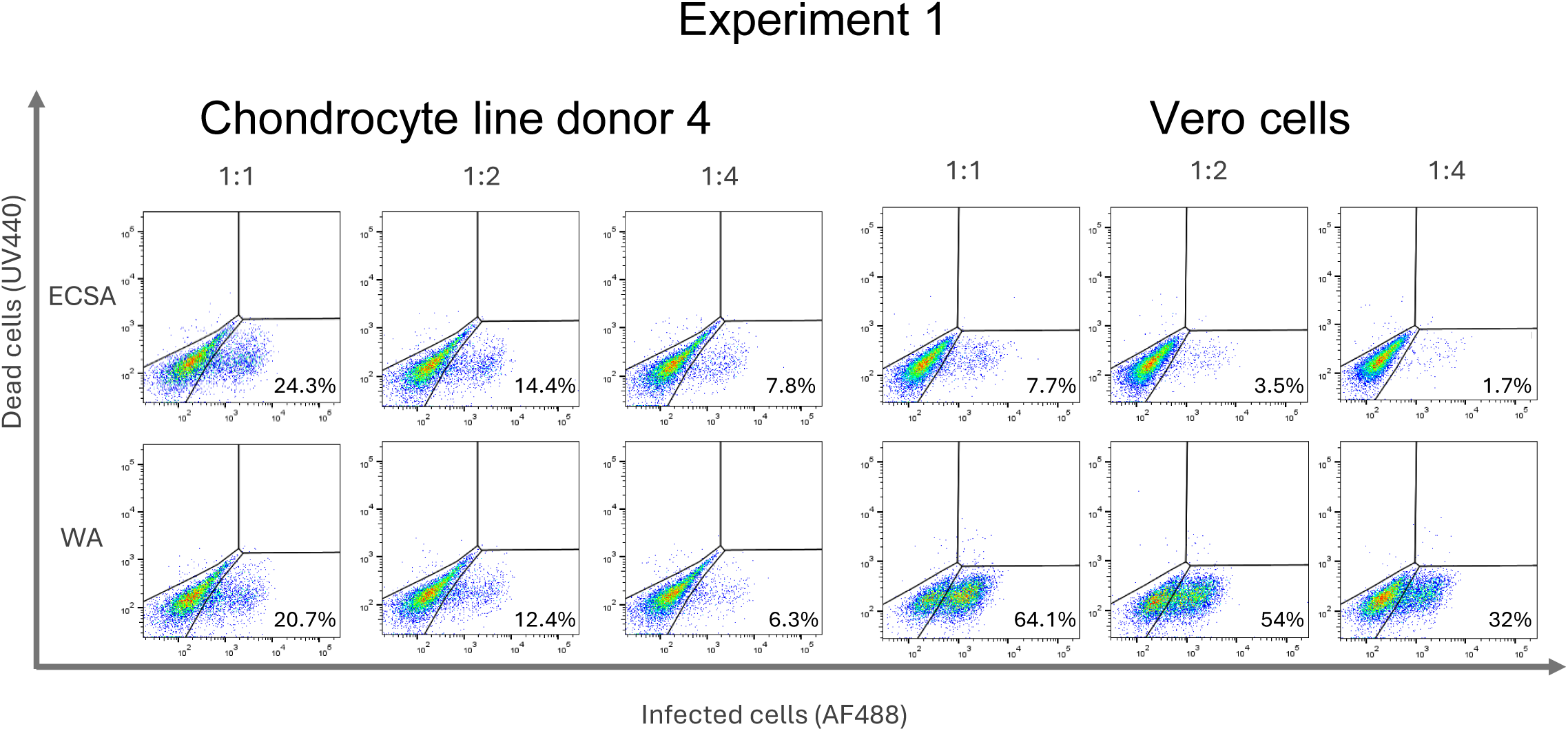
Flow cytometry analysis of CHIKV infection in chondrocytes (donor 0004) and Vero-E6 cells. X-axis is CHIKV infection, y-axis is cell death. Numbers above plots indicate dilution, numbers within plots indicate percentages within each quadrant, with live infected cells bottom right.

**Figure S5:**
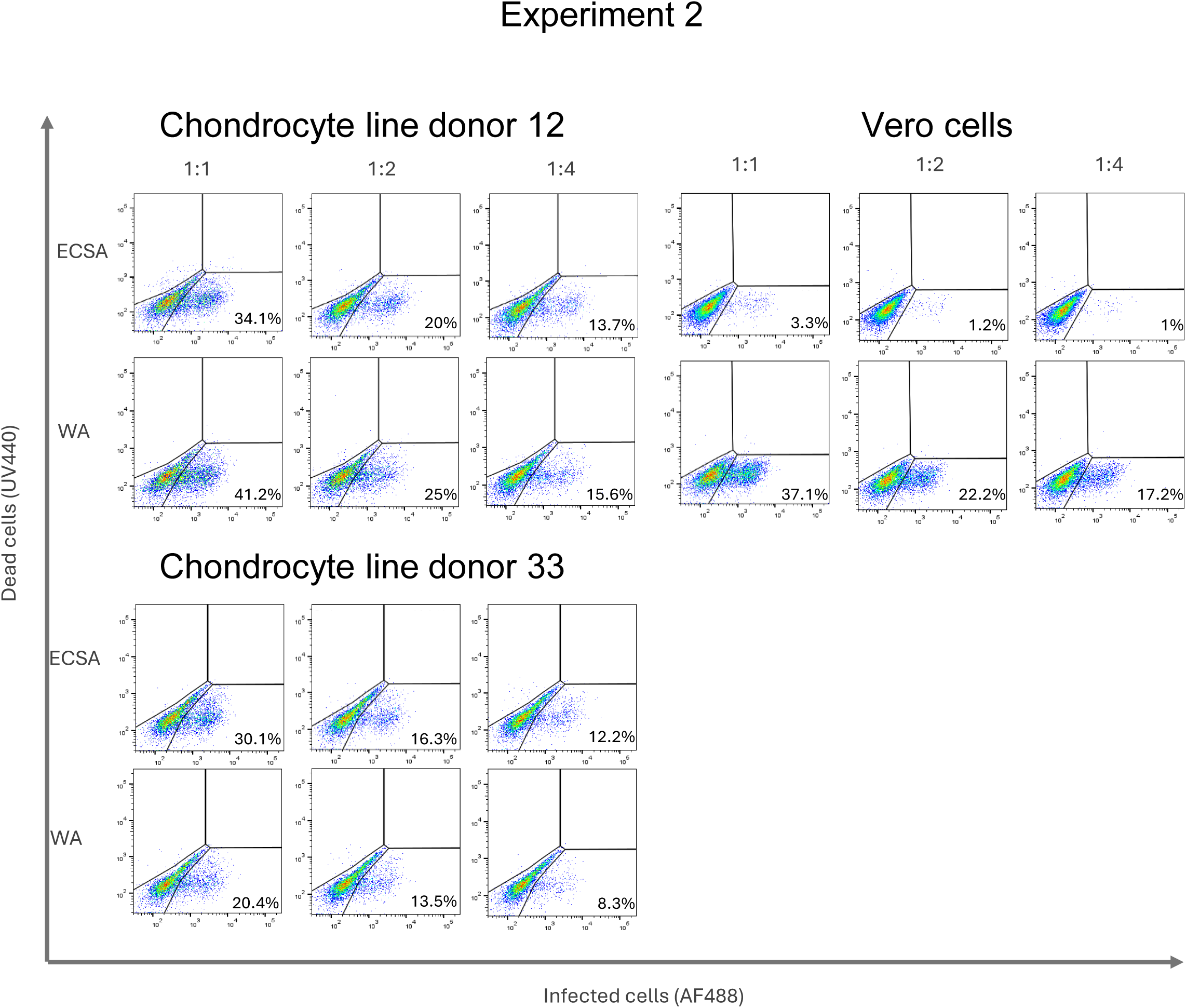
Flow cytometry analysis of CHIKV infection in chondrocytes (donor 0012 and 0033) and Vero-E6 cells. **X-axis is CHIKV infection, y-axis is cell death. Numbers above plots indicate dilution, numbers within plots indicate percentages within each quadrant, with live infected cells bottom right.**

**Figure S6:**
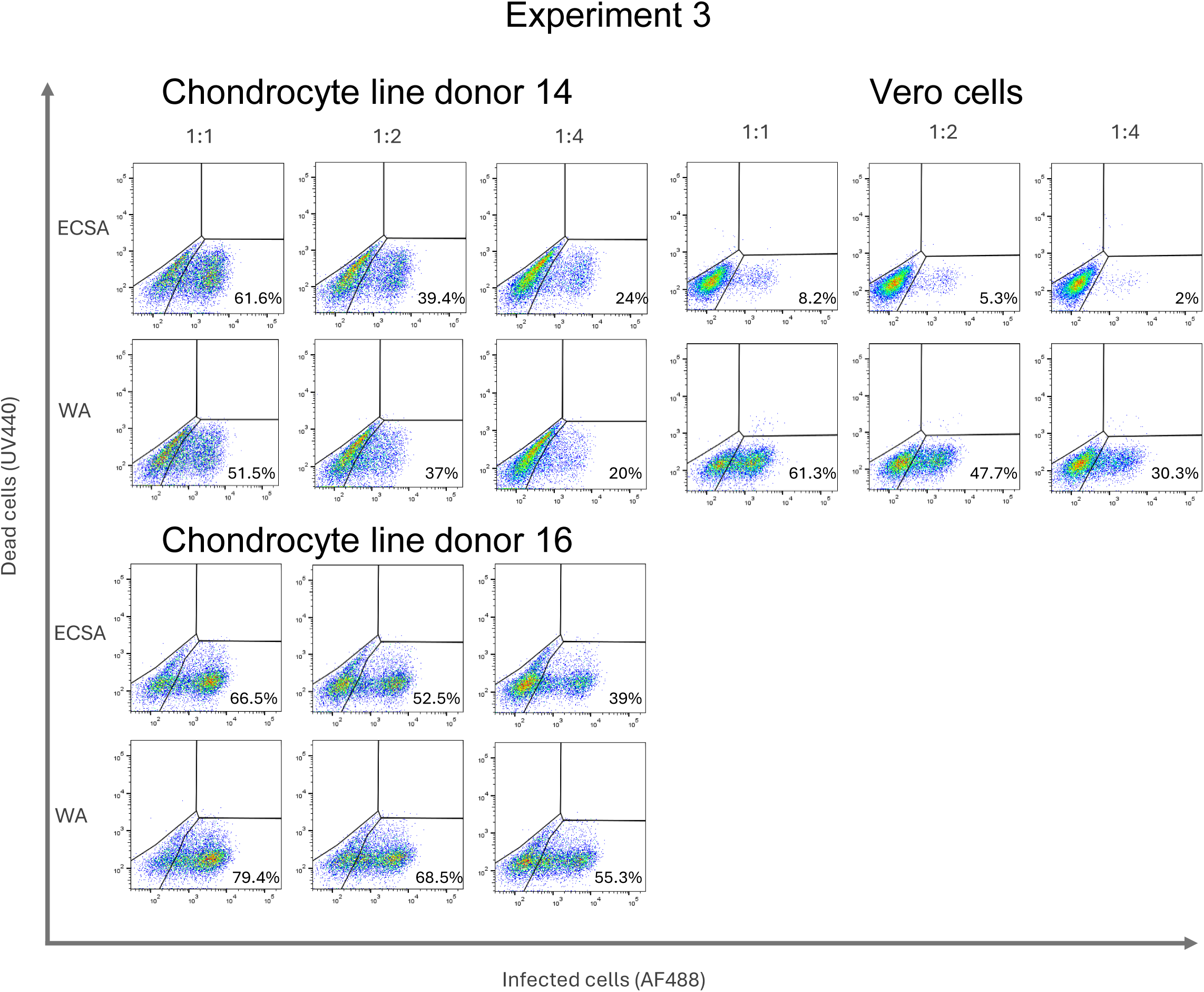
Flow cytometry analysis of CHIKV infection in chondrocytes (donor 0014 and 0016) and Vero-E6 cells. X-axis is CHIKV infection, y-axis is cell death. Numbers above plots indicate dilution, numbers within plots indicate percentages within each quadrant, with live infected cells bottom right.

**Figure S7:**
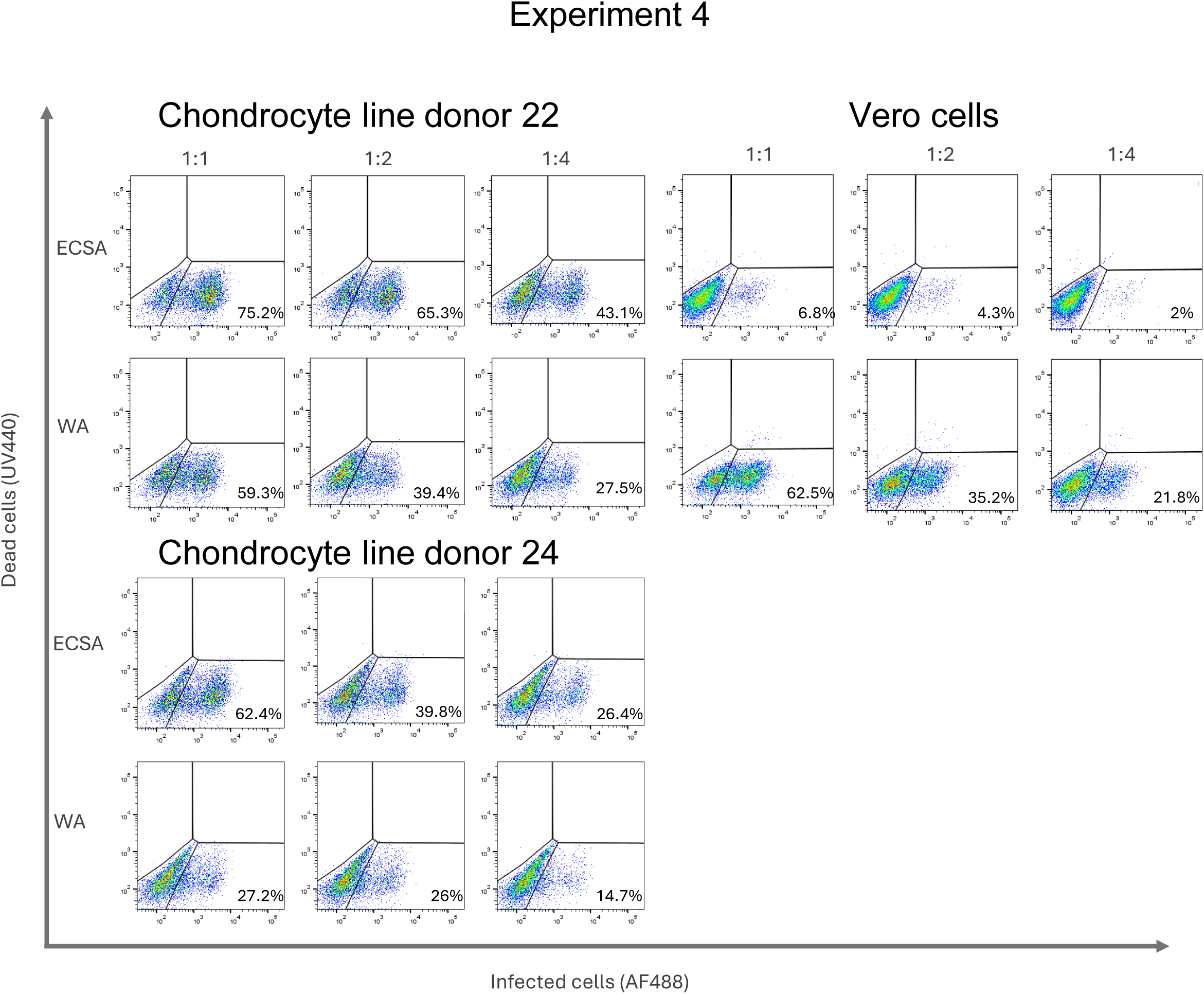
Flow cytometry analysis of CHIKV infection in chondrocytes (donor 0022 and 0024) and Vero-E6 cells. X-axis is CHIKV infection, y-axis is cell death. Numbers above plots indicate dilution, numbers within plots indicate percentages within each quadrant, with live infected cells bottom right.

**Figure S8:**
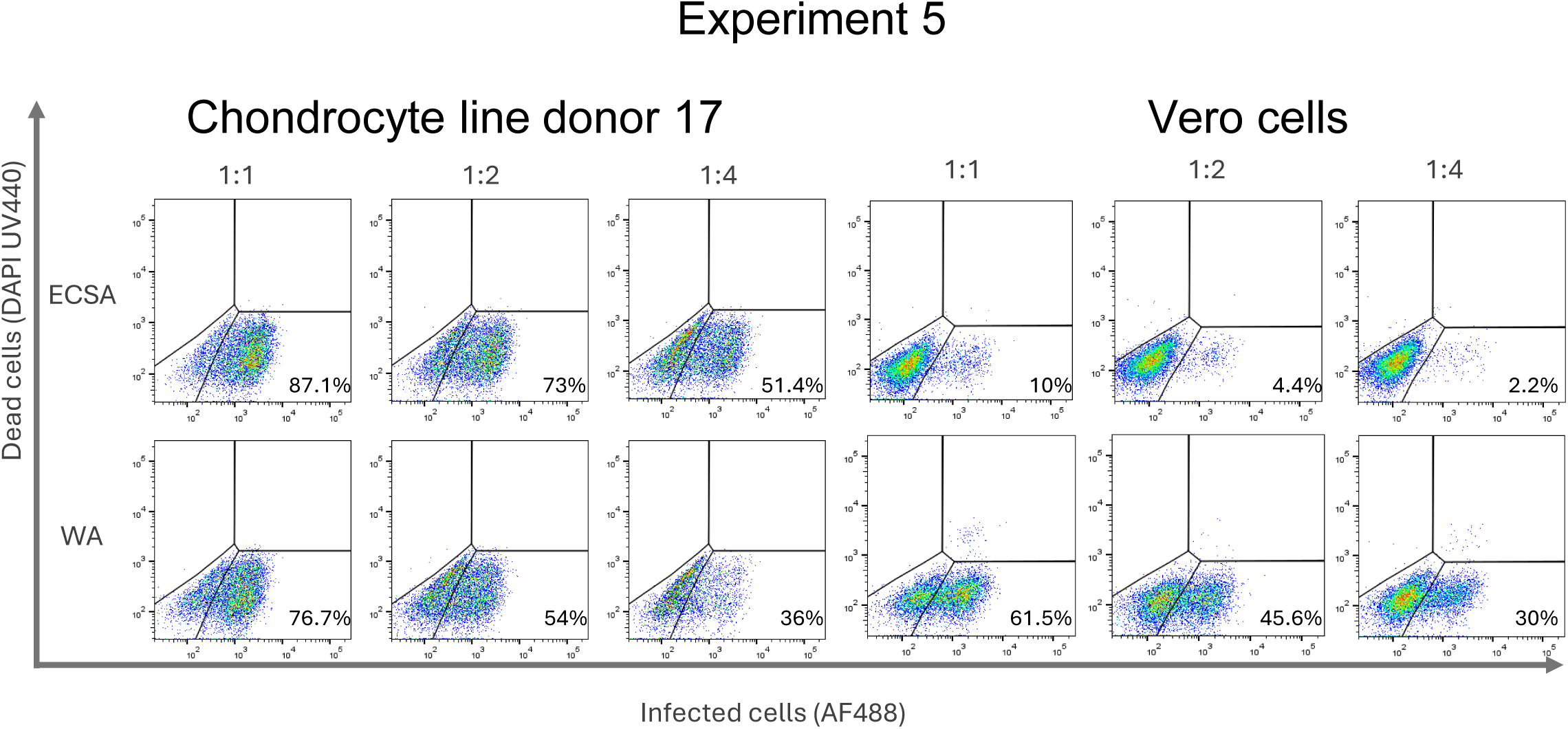
Flow cytometry analysis of CHIKV infection in chondrocytes (donor 0017) and Vero-E6 cells. X-axis is CHIKV infection, y-axis is cell death. Numbers above plots indicate dilution, numbers within plots indicate percentages within each quadrant, with live infected cells bottom right.

**Figure S9:**
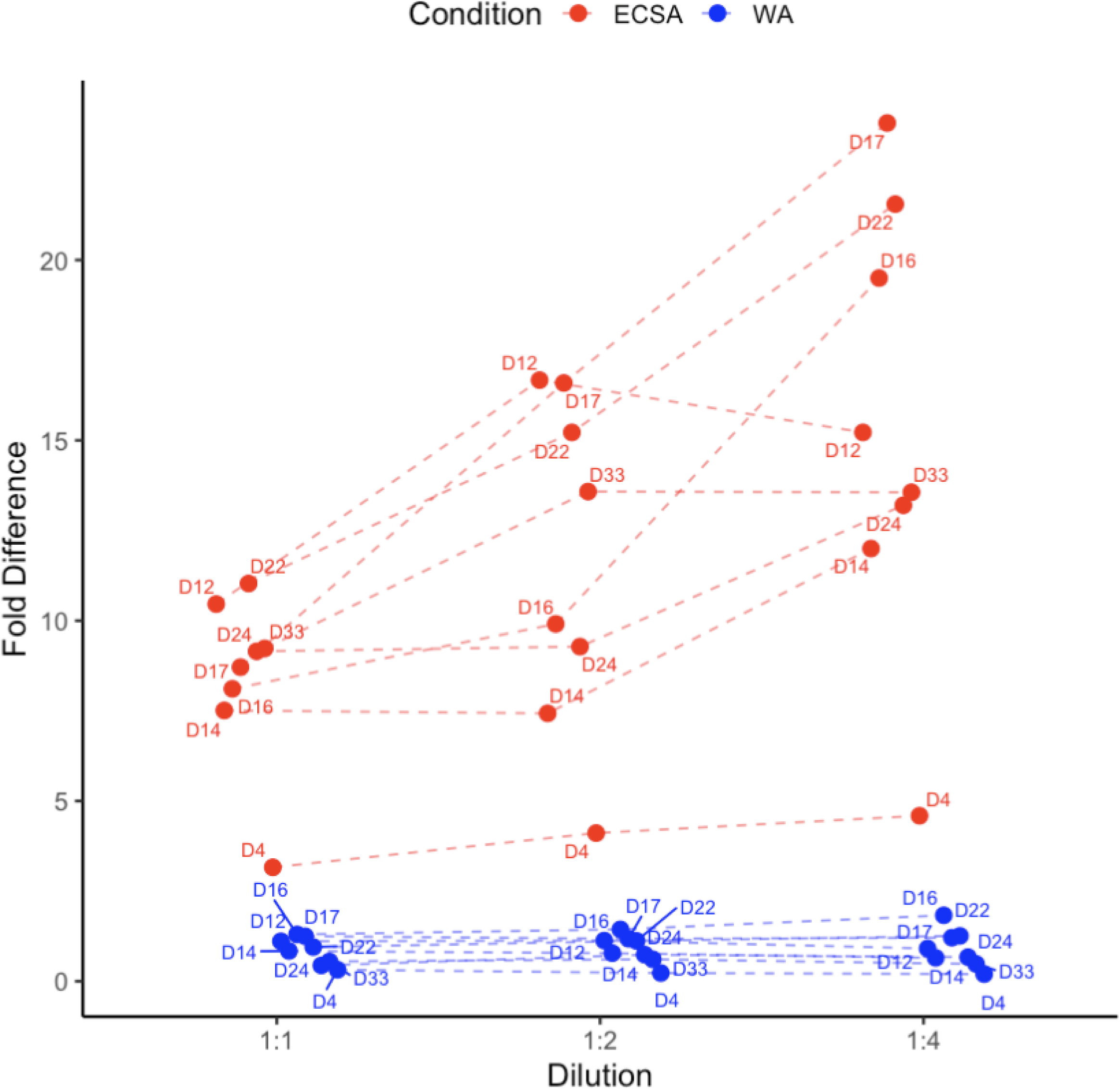
Ratio of CHIKV infection in chondrocytes relative to Vero-E6 cells. Chondrocyte-to-Vero ECSA or WA infection ratios for each donor across three viral dilutions. Individual donors are identified by the number next to each data point.

**Figure S10.**
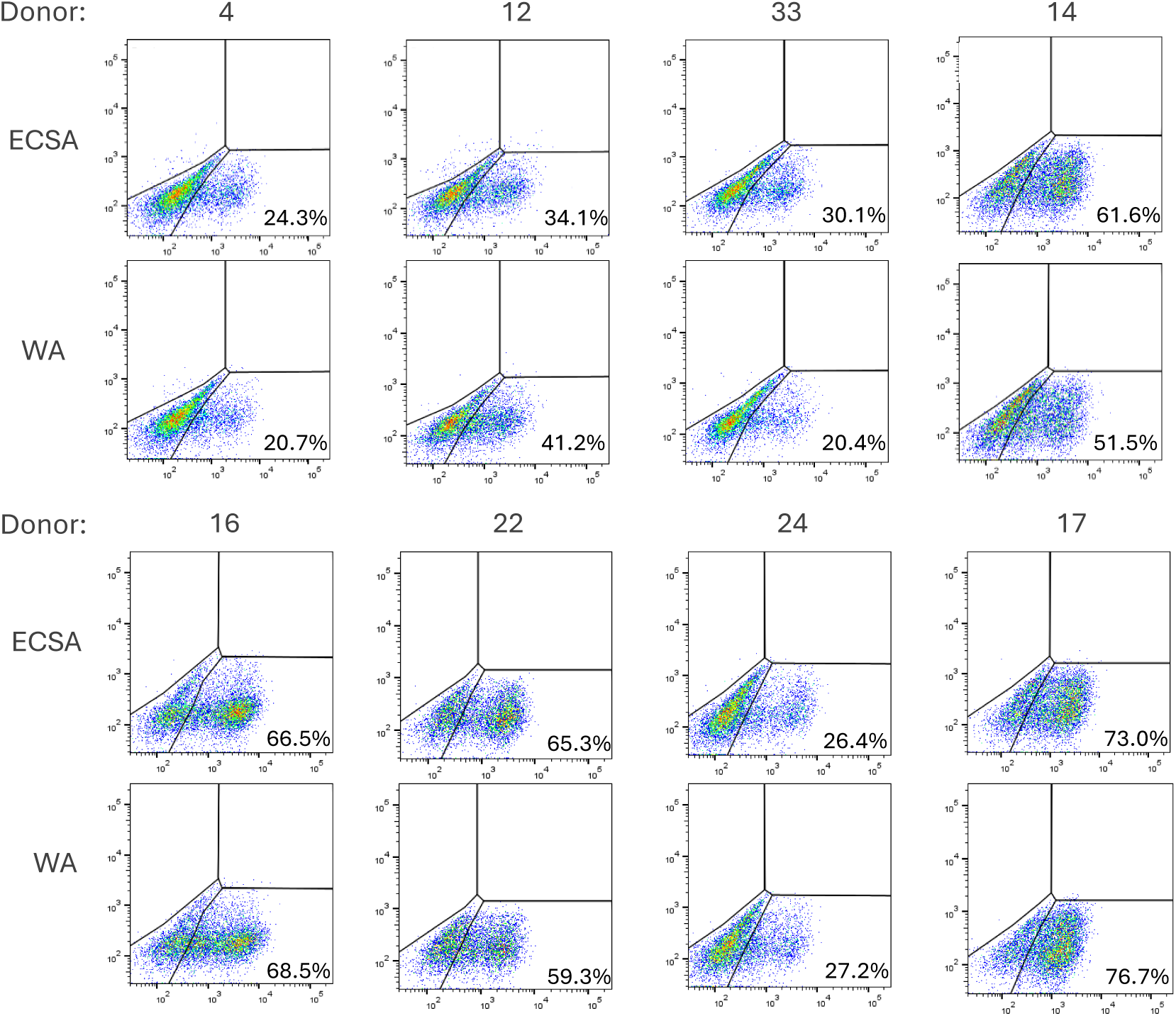
Selection of matched infection conditions for analysis. Flow cytometry-derived infection percentages for ECSA and WA lineages across multiple viral dilutions in primary human chondrocytes from each donor 24 hours post-infection. Dilutions yielding comparable infection levels between lineages (indicated) were selected for downstream RNA-seq analysis. Comparable infection was defined as <10% difference between infection levels in ECSA and WA infected cells.

**Table S1:** Cell type identity verification by transcriptome using SingleR.

|  | Biological replicate |  |  |
| --- | --- | --- | --- |
| Donor number | 1 | 2 | 3 |
| 17 | Chondrocytes | Chondrocytes | Chondrocytes |
| 22 | Chondrocytes | Chondrocytes | Chondrocytes |
| 14 | Chondrocytes | Chondrocytes | Chondrocytes |
| 16 | Chondrocytes | Chondrocytes | Chondrocytes |
| 12 | Chondrocytes | Chondrocytes | Chondrocytes |
| 33 | Chondrocytes | Chondrocytes | Chondrocytes |
| 24 | Chondrocytes | Chondrocytes | Chondrocytes |
| 4 (experiment 1) | MSC | MSC | MSC* |
| 4 (experiment 2) | MSC | MSC | MSC |
| 8 | Chondrocytes | Chondrocytes | Chondrocytes* |
| 1 | MSC | Fibroblast | Fibroblast * |

SingleR reference-based classification was performed using the Human Primary Cell Atlas as the reference dataset. \*RNA-Seq performed in the presence of infection in this replicate.

**Table S2:** Top principal component 1 loading genes.

| Ensembl id | Gene name | PC1 contribution | Function (GeneCards) |
| --- | --- | --- | --- |
| **ENSG00000157601** | MX1 | 0.091 | Interferon-induced antiviral protein |
| **ENSG00000185745** | IFIT1 | 0.072 | Interferon-induced antiviral protein |
| **ENSG00000130518** | IQCN | 0.069 | Encodes a protein that contains IQ motifs (canonical calmodulin-binding domains involved in calcium signaling) |
| **ENSG00000119922** | IFIT2 | 0.067 | Interferon-induced antiviral protein |
| **ENSG00000135114** | OASL | 0.064 | Interferon-induced antiviral protein |
| **ENSG00000142871** | CCN1 | -0.056 | Promotes cell proliferation, chemotaxis, angiogenesis and cell adhesion, and down-regulates the expression of alpha-1 and alpha-2 subunits of collagen type-1 |
| **ENSG00000130589** | HELZ2 | 0.054 | Nuclear zinc finger helicase that acts as a transcriptional co-activator of PPARα which can suppress inflammatory signaling |
| **ENSG00000118523** | CCN2 | -0.054 | Promotes chondrocyte differentiation |
| **ENSG00000187608** | ISG15 | 0.054 | Interferon-induced antiviral protein |
| **ENSG00000111335** | OAS2 | 0.053 | Interferon-induced antiviral protein |

**Table S3:** Top principal component 2 loading genes.

| Ensembl id | Gene name | PC2 contribution | Function |
| --- | --- | --- | --- |
| **ENSG00000174327** | SLC16A13 | 0.18 | Predicted to be involved in monocarboxylic acid transport and transmembrane transport; possible export of lactate in glycolysis |
| **ENSG00000171435** | KSR2 | 0.15 | Membrane-associated serine/threonine kinase involved in Ras and calcium-mediated signaling and negative regulator of MAP3K3-mediated activation of ERK, JNK and NFkB pathways |
| **ENSG00000224586** | GPX5 | 0.15 | Protects cells from oxidative damage |
| **ENSG00000176165** | FOXG1 | 0.14 | Transcriptional regulator controlling cell proliferation, survival, and differentiation |
| **ENSG00000181449** | SOX2 | 0.12 | One of four Yamanaka factors that when combined, are sufficient to reprogram differentiated cells to an embryonic-like state |
| **ENSG00000134249** | ADAM30 | 0.12 | ADAM-family metalloprotease implicated in cell-cell and cell-matrix interactions |
| **ENSG00000157601** | MX1 | 0.11 | Interferon-induced antiviral protein |
| **ENSG00000148400** | NOTCH1 | 0.11 | Can protect joints or exacerbate joint injury depending on signal strength/duration/context (see also Zhaoyang Liu et al., Sci. Signal.8,ra71-ra71(2015).DOI:10.1126/scisignal.aaa3792) |
| **ENSG00000204970** | PCDHA1 | 0.10 | Potential calcium-dependent cell-adhesion protein |
| **ENSG00000126218** | F10 | 0.10 | Coagulation factor which triggers the production of pro-inflammatory cytokines |

